# Comparison of RNA extraction methods for the detection of SARS-CoV-2 by RT-PCR

**DOI:** 10.1101/2020.08.13.20172494

**Authors:** Ossia M. Eichhoff, Elisa Bellini, Reto Lienhard, Wendelin J. Stark, Philippe Bechtold, Robert N. Grass, Philipp P. Bosshard, Mitchell P. Levesque

**Affiliations:** Department of Dermatology, University Hospital Zurich, 8091 Zurich, Switzerland Switzerland; Faculty of Medicine, University of Zurich, 8006 Zurich, Switzerland; ADMED Microbiologie, 2300 La Chaux-de-Fonds, Switzerland; Department of Chemical and Applied Biosciences, ETH Zurich, 8093 Zurich, Switzerland

## Abstract

**Objectives:** The SARS-CoV-2 pandemic outbreak has stressed health care systems as well as medical supply chains, but diagnostic testing is an essential public health measure to control viral spread. Here we test the suitability of different RNA extraction methods for integration into a diagnostic workflow for coronavirus testing.

**Methods:** We applied six RNA extraction methods on the same 24 SARS-CoV-2 positive patient samples and quantified their results by subsequent reverse-transcriptase PCR (RT-PCR) of three viral genes. These methods included a) column-based extraction, b) phenol-chloroform extraction, as well as c) extraction using magnetic beads (i.e., one commercial kit as well as three different magnetic beads in combination with home-brewed buffers and solutions).

**Results:** We achieved diagnostic-quality RT-PCR results with all methods, and there was no significant difference between the tested methods, except for one magnetic bead protocol with home-brewed buffers, in which the number of positive tested genes was significantly lower.

**Conclusions:** Five of the six RNA extraction methods are interchangeable in a diagnostic workflow. Since some methods are more scalable than others, and have comparable results on RT-PCR quantitation, they may be more amenable to high-throughput sample processing pipelines.

**Graphical abstract:** 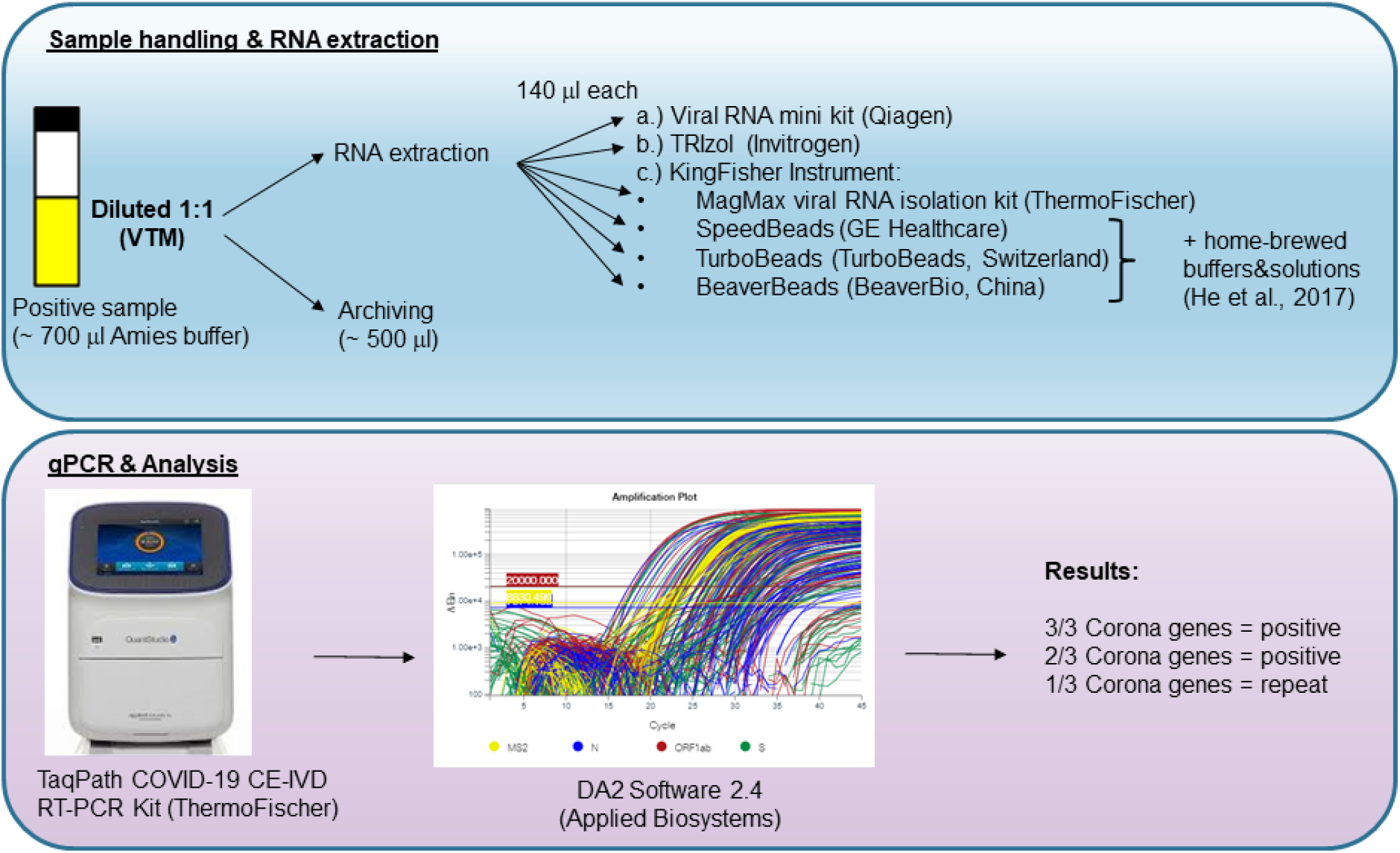

## Introduction

With over 4 million confirmed active cases worldwide, the coronavirus pandemic is a historical outbreak with enormous consequences for national health care systems and economies. On December 31st 2019, the World Health Organization (WHO) was informed about the detection of a cluster of cases of pneumonia with unknown origin appearing in Wuhan, Hubei Province of China [1]. Investigation of patients suffering from this new respiratory disease revealed that the cause was a novel coronavirus, now named severe acute respiratory syndrome coronavirus 2 (SARS-CoV-2) which caused the coronavirus disease 2019 (COVID-19) [2, 3]. In a short period, SARS-CoV-2 spread to a dozen countries and within a few months developed into a pandemic outbreak [4]. Until an efficacious and safe vaccine is available, the only way to prevent further spread of the virus is to dramatically reduce infection rates. The WHO recommends several public health measures: (i) rapid diagnosis and immediate isolation of cases, (ii) rigorous tracking, and (iii) precautionary self-isolation [5]. These strategies also mean that testing must be widely available and the barriers to testing have to be as low as possible. Thus, hundreds of thousands of tests need to be available daily worldwide, which challenges global supply chains and the production of reagents necessary for diagnostic testing. In order to reduce supply chain vulnerabilities and limit dependencies on single suppliers, we compared different RNA extraction protocols to establish our coronavirus diagnostics workflow at the Department of Dermatology, University Hospital of Zurich, which could be used for subsequent detection of viral RNA by reverse-transcriptase quantitative PCR (RT-PCR).

## Methods

### Biological specimens

We received 24 nasal or mucosal swabs from confirmed SARS-CoV-2 positive patients from the ADMED laboratory, Switzerland.

### RNA extraction

Nasal or mucosal swabs were provided in Amies medium and were diluted 1:1 with viral transport medium (VTM prepared according to the protocol of the Center of disease control (CDC): https://www.cdc.gov/coronavirus/2019-ncov/downloads/Viral-Transport-Medium.pdf). For each RNA extraction method an aliquot of 140 μl was taken.

a. The viral RNA mini kit (Qiagen, Switzerland) was used according to the manufacturer’s protocol. Briefly, 140 μl were mixed with 560 μl AVL-carrier RNA buffer and incubated for 10 minutes at room temperature. Column-based RNA extraction was carried out using the Qiacube automated column extraction device from Qiagen. The elution volume was set to 100 μl.
b. TRIzol-based phenol-chloroform RNA extraction (Invitrogen, Thermo Fisher, Switzerland) was carried out according to the manufacturer’s protocol and 140 μl sample was mixed into 860 μl of TRIzol solution. RNA was dissolved in 100 μl DEPC-treated water.
c. For the comparison of different magnetic bead-based protocols, we used SpeedBeads (GE Healthcare, USA), BeaverBeads (BeaverBio, China), TurboBeads (TurboBeads, Switzerland), and the magnetic bead-based extraction kit for the KingFisher instrument (MagMax, Thermo Fischer). The MagMax isolation kit was used according to the manufacturer’s protocol with a sample input of 140 and the instrument software provided on the Thermo Fischer webpage. Magnetic bead-based extraction using the KingFisher™ Flex Purification System (Thermo Fisher) together with home-brewed buffers and solutions either with SpeedBeads, BeaverBeads or TurboBeads was based on the publication from He et al, 2017 [6]. Specifically, a total of 200 μl diluted sample (140 μl patient sample + 60 μl VTM) was incubated for 10 minutes at room-temperature in a 96-DeepWell plate together with 300 μl of lysis buffer (2 M guanidinium thiocyanate, 80 mM dithiothreitol, 25 mM sodium citrate, 20 μg/ml glycogen and 0.5% Triton-X 100, pH 6) containing 1 μg carrier-RNA. The extraction protocol was started on the KingFisher instrument with the following steps 1) heating of the plate at 80°C for 10 minutes and cooling back to room temperature (RT), 2) instrument paused while adding 480 μl of 100% EtOH and 20 μl magnetic beads to each well of the 96-DeepWell plate, 3) program continues with a 5 minute incubation at room temperature which allows the nucleic acids to be absorbed by the magnetic beads, 4) the instrument will collect the beads and wash them twice in 70% EtOH. Nucleic acids were eluted in the elution plate containing 100 μl of DEPC-treated water for all KingFisher methods.

### RT-PCR

The detection of viral RNA in each sample was performed via RT-PCR using the TaqPath COVID-19 CE-IVD RT-PCR Kit, according to the manufacturer’s protocol with a RNA sample input of 10 μl. The MS2 internal performance control was added to the PCR master mix. The RT-PCR was run on a QuantStudio 5 real-time PCR-System (Applied Biosystems, Switzerland) and data were analyzed with the Design and Analysis Software DA 2.4 (Applied Biosystems).

## Results & Discussion

We compared the feasibility of different commercially available and home-brewed RNA isolation techniques for a SARS-CoV-2 diagnostics workflow with the aim to test whether these techniques can be inter-changeable in crises when supply-chains are unreliable. We chose to evaluate the RNA extraction efficiency of each method via the multiplex RT-PCR TaqPath COVID-19 CE-IVD RT-PCR Kit, which was approved as a diagnostic tool by the FDA and as a CE mark throughout Europe. We found that all methods tested here, except using magnetic-beads produced by BeaverBio, could be used to make diagnostic-quality RNA extractions, as there was no statistically significant difference between the results as tested with Fisher’s exact tests. Using BeaverBeads, however, required two samples to be repeated due to inconclusive results (e.g., 2 out of 3 genes negative, Figure 1 +2) and the overall number of positive tested genes was significantly lower as compared to the TRIzol method (64/72 versus 72/72; p = 0.0064), which detected all three genes in each sample. All other methods did not significantly differ from TRIzol (p > 0.5), but in all other methods single genes were not recognized, increasing the risk of false negative results. Importantly, methods that require intensive manual pipetting (TRIzol) or allow only low-scale throughput (Qiacube, 12 samples/run) are not optimal for a daily routine process with 200+ samples and therefore magnetic-bead based extraction using a KingFisher instrument is advantageous, since it processes 96 samples/run. In conclusion, two of the three magnetic beads with home-brewed buffers and solutions can be equally used in comparison to the MagMax manufacturer’s kit. Thus, the dependency on suppliers in times of crisis can be decreased by implementing comparable techniques for the isolation of viral RNA in a laboratory diagnostic workflow.

**Fig 1:**
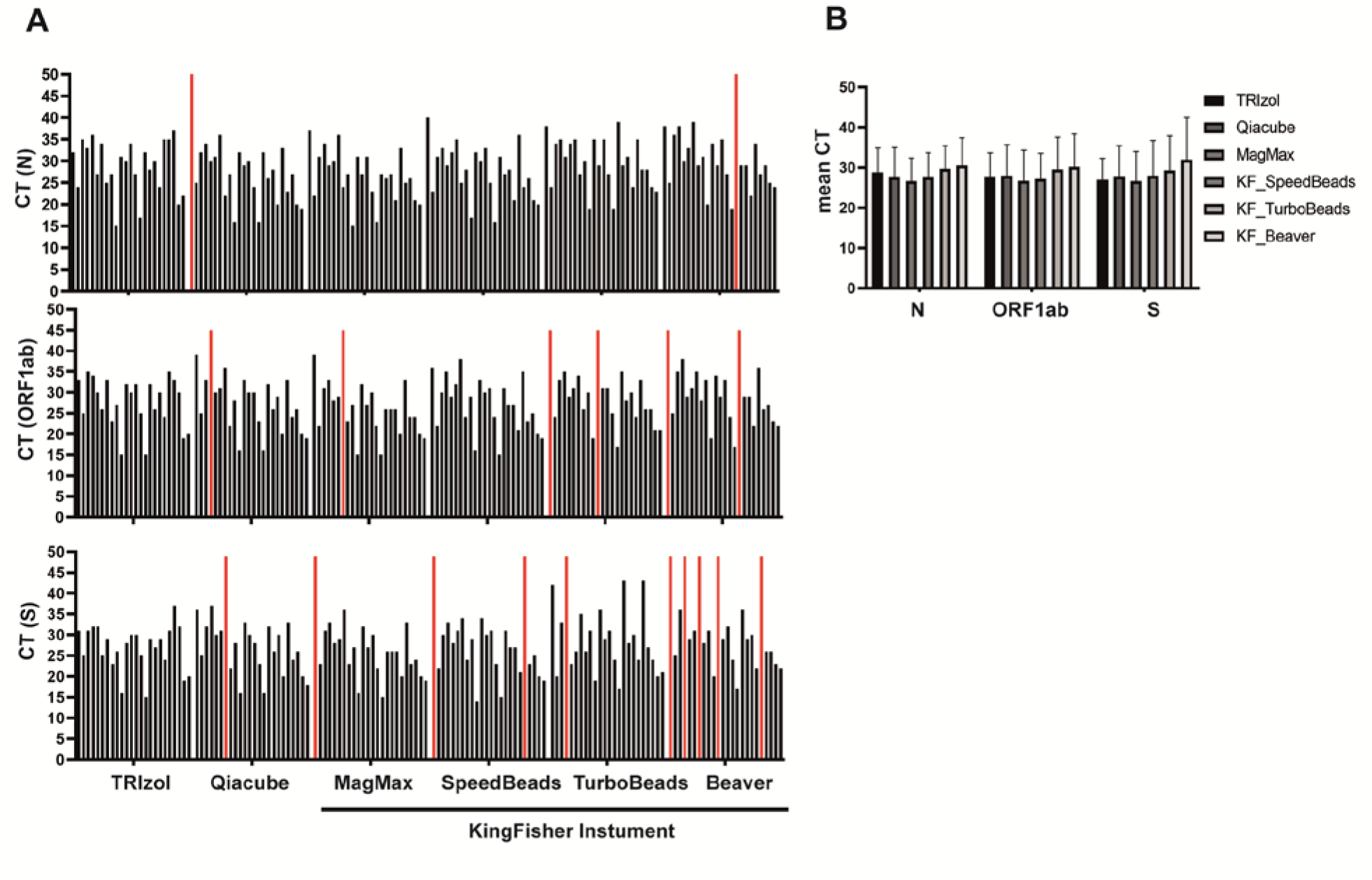
(A) Summary of the RT-PCR results comparing CT values for three SARS-CoV-2 specific sequences (N, ORF1ab and S). Each bar represents one patient sample, and CT values with undetermined amplification are set to a CT of 50 and are shown in red. (B) Comparison of the mean CT values for each extraction method and each coronavirus target gene.

**Fig 2:**
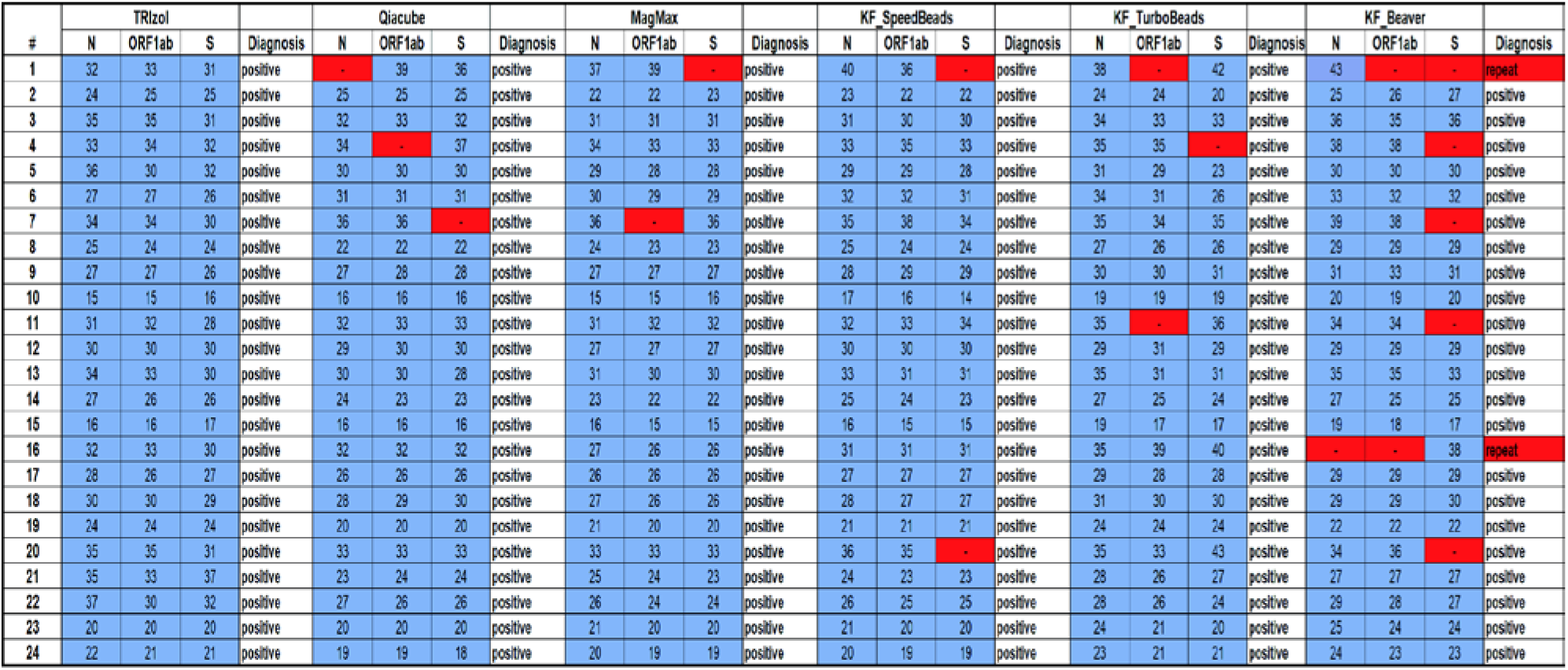
Summary table of the CT values for each patient and extraction method including the diagnostic result. Undetermined CT values are highlighted in red (-).

## Data Availability

All data generated for this manuscript are included in figures and tables

